# How to Demonstrate the Glucose Specificity of a Non-Invasive CGM: A Case Study of the SKAMo-2 Clinical Trial and Neogly™

**DOI:** 10.64898/2026.08.17.26360581

**Authors:** Romain Blanc, Pierre Blandin, Jean-Guillaume Coutard, Kevin Jourde, Hélène Marie, Pierre-Yves Benhamou

## Abstract

**Background:** Every non-invasive continuous glucose monitoring (NI-CGM) technology introduced into the landscape faces the same skeptical question, from regulators, clinicians, and competing developers alike: is the candidate signal actually specific to glucose, or does an apparently reasonable accuracy figure simply reflect a model fitting to motion, temperature, calibration offset, or trial-duration artifact? Existing evaluation practice does not answer this question directly. NI-CGM performance is instead reported almost exclusively with metrics inherited from minimally invasive, subcutaneous CGM, the Mean Absolute Relative Difference (MARD), Clarke/Parkes error grids, and ISO 15197-style agreement rates, which were designed for sensors whose glucose specificity is already chemically established and which therefore take specificity as a premise rather than treating it as a result to be demonstrated.

**Methods:** We present a methodology for demonstrating NI-CGM technology glucose specificity during the algorithm-development phase, and illustrate it with a case study based on a quantum-cascade-laser (QCL) photoacoustic NI-CGM device (Neogly™) evaluated in the SKAMo-2 free-living clinical trial (eight participants with type 1 diabetes). The methodology combines a white-noise control, a constant-glycemia control, a sensor-ablation control that removes the candidate physical signal while retaining auxiliary covariates, and explicit reporting of the train/test generalization level, so that a reported MARD can be read as evidence of specificity rather than taken on faith.

**Results:** Removing the mid-infrared photoacoustic (PA) signal from the model while retaining all auxiliary sensors (accelerometer, skin temperature, hygrometry, PPG) degraded performance at every generalization level tested, inter-patient MARD rose from 35.0% with the PA signal to 43.1% without it, and intra-experimentation MARD rose from 22.5% to 23.9%, providing direct, internal evidence that the PA channel itself, and not merely the auxiliary covariates, carries glucose-specific information. At the same time, an algorithm trained on pure Gaussian noise produced a MARD of 25% over short test windows, and a trivial constant-glycemia predictor outperformed every machine-learning model tested when generalization was extended from a single recording to an unseen patient (MARD 55% for the naive constant model versus 37% for a deep neural network on inter-patient splits). Reported in isolation, any of these MARD values is uninterpretable; reported against one another, they jointly demonstrate that the signal is specific to glucose while also bounding how much of the headline accuracy figure that specificity currently explains.

**Conclusions:** We propose a specificity-demonstration methodology for NI-CGM technology development, comprising (1) signal quality gating prior to any algorithm benchmarking, (2) a white-noise control to test for genuine information content, (3) a constant-glycemia control to expose trial-duration bias, (4) a sensor-ablation control that isolates the contribution of the candidate physical signal from auxiliary covariates, (5) explicit reporting of the data-splitting generalization level (intra-experimentation, intra-patient, inter-patient). This methodology answers a question that precedes clinical accuracy reporting and that recognized clinical frameworks such as the IFCC Working Group on CGM’s Dynamic Glucose Regions guideline are not designed to answer: not how accurate is the device, but is the device measuring glucose at all. We argue that without these controls, MARD and error-grid values for NI-CGM are not comparable across studies and may either overstate clinical readiness or undermine promising technologies. We recommend that this specificity methodology be applied routinely once a candidate NI-CGM sensor reaches algorithm-development stage, alongside and as a deliberate complement to IFCC-style clinical accuracy reporting once the device is mature enough for that evaluation.

## 1. Introduction

Subcutaneous, minimally invasive continuous glucose monitoring (CGM) has become a standard of care for insulin-treated diabetes, and the metrics used to evaluate it, chiefly the Mean Absolute Relative Difference (MARD) and the Clarke or Parkes error grids, are now familiar to the entire diabetes technology community.^1,2^ These metrics were developed, refined, and validated over nearly two decades against electrochemical enzymatic sensors whose underlying signal-to-glucose relationship is well characterized and whose principal sources of error are calibration drift, sensor warm-up, and the physiological lag between interstitial and capillary glucose.^3^

A new generation of non-invasive CGM (NI-CGM) devices, based on infrared optical absorption, photoacoustic spectroscopy, bioimpedance, radiofrequency/microwave sensing, Raman spectroscopy or photoplethysmography-derived surrogates, is now being evaluated with the same toolbox.^4, 5, 6, 7, 8^ This is a reasonable starting point, since regulatory bodies and clinicians need a common language to compare devices. However, NI-CGM systems differ from their subcutaneous counterparts in a way that legacy metrics do not capture: the existence of a glucose-specific, extractable signal is itself the object of investigation, not a settled premise. Furthermore, the difficulty of developing wearable products based on these new technologies, reproducible at scale, limits drastically the implementation of generalizable predictive algorithms.

Finally, in practice, the question every team developing a non-invasive sensor is eventually asked, by reviewers, by investors, by competitors, and by clinically skeptical audiences: is this actually measuring glucose? An electrochemical sensor’s MARD describes how accurately a known signal is converted into a glucose value. For many NI-CGM modalities, a high or low MARD can instead simply reflect how much of the test window happened to contain stable glycemia, how the algorithm was calibrated, or how the reference device’s own error propagated into the comparison independently of whether the underlying physical signal contains any glucose information.^9, 10^

This distinction matters because MARD, by construction, rewards any predictor that stays close to the reference value, including predictors with no physiological basis whatsoever. A model that simply repeats the calibration value for the rest of a short recording, or a model trained on synthetic noise matched to the empirical mean and variance of the cohort’s glucose levels, can both achieve MARD values in the same range historically considered “acceptable” for CGM, not because they are accurate, but because real-world glycemic excursions over short windows are frequently smaller than the errors tolerated by the metric.^9,11^ Several scoping reviews of CGM clinical evaluations have already noted the extreme heterogeneity in trial design, population, reference device, and pairing method that undermines cross-study comparison of MARD values even for mature, subcutaneous sensors^12^. For NI-CGM, where the signal itself is contested, this heterogeneity becomes a more serious problem because it can mask the absence of a genuine glycemic signal behind an apparently reasonable accuracy figure.

In this article, we propose a methodology for demonstrating the glucose specificity of an NI-CGM signal during the algorithm-development phase, that is, for answering the prior, narrower question of whether a candidate signal measures glucose at all, before asking how accurately it does so. This methodology is intended to complement, not replace, the clinical accuracy metrics required for regulatory submission, including the framework proposed by IFCC Working Group on CGM’s^28^. Such metrics are designed to be applied once a sensor and algorithm are already working, to characterize accuracy across clinically meaningful glucose ranges. The methodology proposed here is designed to be applied earlier, to establish that there is a working, glucose-specific signal to characterize in the first place. We argue that this specificity question deserves its own explicit methodology rather than being left implicit in a single accuracy number. Once a sensor has cleared this bar, the IFCC approach is the right tool for the clinical-accuracy evaluation that should follow. The methodology adds explicit negative controls (a white-noise benchmark and a constant-glycemia benchmark), a sensor-ablation control that isolates the contribution of the candidate physical signal from auxiliary covariates, a structured reporting of the generalization level used to split training and test data. We illustrate the methodology with a case study based on Neogly™, a wearable mid-infrared quantum-cascade-laser (QCL) photoacoustic NI-CGM device, evaluated during the SKAMo-2 free-living clinical trial on eight participants with type 1 diabetes.^13^

## 2. The Current CGM Evaluation Toolbox and Its Limits for NI-CGM

### 2.1 Point accuracy: MARD

MARD is calculated as the mean of the absolute relative differences between paired sensor and reference glucose values, expressed as a percentage (Equation 1).^2,14^

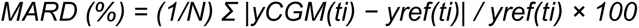

with yCGM(ti) the glucose value measured at the i^th^ time stamp ti of the acquisition, by the evaluated system, yref(ti) the glucose value measured at the same time with the reference system, and N the total number of time stamps and consequently values for each system. MARD is an arithmetic average computed across all sensors, participants, and glycemic ranges over a sensor’s wear life.^14^ A low aggregate MARD can therefore mask localized failures, a sensor that performs poorly during the first hours of wear, or in the hypoglycemic range. It may still report an acceptable overall value if the rest of the recording is stable.^14,15^ MARD is also not a pure descriptor of the device: it is influenced by the reference method’s own error, which propagates directly into the calculated value and acts as a positive offset. If a study uses a domestic capillary meter (typically 6–10% error) instead of a laboratory-grade analyzer, the comparison partly evaluates the reference device rather than the sensor under test.^14,16^ Finally, MARD is a static, point-in-time metric: it does not credit or penalize a sensor’s ability to track the direction and rate of glycemic change, which is clinically important for detecting sudden excursions, hypoglycemia avoidance, and for closed-loop insulin delivery.^14,17^

### 2.2 Agreement rates and ISO/FDA thresholds

Agreement rates (ARs) express the percentage of paired readings within a fixed numerical tolerance, typically ±15 mg/dL below 100 mg/dL and ±15% above 100 mg/dL, following the structure of ISO 15197:2013 for self-monitoring blood glucose meters, and adapted by several groups to CGM and NI-CGM reporting.^18,19^ The FDA’s integrated CGM (iCGM) special controls (21 CFR 862.1355) extend this logic with explicit, range-specific accuracy and concurrence requirements (for example, agreement within ±15 mg/dL or ±15% across defined glucose bands, plus zonal constraints preventing iCGM values below 70 mg/dL from corresponding to a reference reading above 180 mg/dL, and vice versa), evaluated as lower one-sided 95% confidence bounds rather than simple point estimates.^20,21^ Agreement rates and iCGM-style bounds are an improvement over a single aggregate MARD because they expose performance separately in clinically critical ranges. However they remain point-in-time comparisons, and inherit the same dependency on trial design and reference accuracy described above.

### 2.3 Clinical risk: the Clarke and Parkes error grids

The Clarke Error Grid Analysis (EGA), introduced in 1987, plots reference glucose on the x-axis against sensor glucose on the y-axis. The plane is divided into five zones (A through E) according to the clinical consequence of acting on the paired values. Zone A (clinically accurate) and Zone B (benign error) are considered acceptable; Zones C, D, and E represent overcorrection, dangerous failure to detect a glycemic extreme, and erroneous treatment, respectively, and should each be close to 0% of paired data.^1,22^ Regulatory practice typically demands that more than 99% of paired points fall in Zones A and B combined^.1^ The Parkes (Consensus) Error Grid refines zone boundaries based on clinician consensus on treatment risk and is used interchangeably with the Clarke grid in much of the CGM literature.^4,23^ Like MARD, however, both grids evaluate each point independently and therefore share its principal weakness: a sensor that reports a flat or lagging trace can still land predominantly in Zone A during periods of stable glycemia, even if it would fail dynamically during a rapid excursion.^17,24^

### 2.4 Diagnostic and dynamic extensions

Several extensions address the static, point-in-time nature of MARD and the error grids. Sensitivity and specificity for hypoglycemia or hyperglycemia detection, summarized via ROC curves and pooled across studies using bivariate random-effects meta-analysis, evaluate a sensor’s diagnostic performance rather than its numerical accuracy.^25, 26^ Continuous Glucose–Error Grid Analysis (CG-EGA), introduced by Kovatchev and colleagues, decomposes accuracy into a point-error grid (P-EGA) and a rate-error grid (R-EGA), so that a sensor’s ability to track the direction and speed of glycemic change is scored independently of its point accuracy.^17,24^ Continuous Glucose Deviation Interval and Variability Analysis (CG-DIVA) uses tolerance intervals to describe the distribution of deviations and between-sensor variability, with particular attention to the incidence of large errors rather than only the mean error.^27^ Bootstrap-based bias-corrected and accelerated (BCa) confidence intervals on agreement rates, which account for the clustered, repeated-measures structure of CGM data within a participant, have been proposed as a more defensible basis for FDA-style accuracy claims than naive confidence intervals that treat every paired point as independent.^28^ The Diabetes Technology Society (DTS) Error Grid replaces the discontinuous boundaries of the Clarke and Parkes grids with smooth, straight-line zones common to both glucose meters and CGM, and quantifies the relationship between Zone A occupancy and MARD (approximately a 0.33-percentage-point MARD change per percentage-point change in Zone A).^29^ More recently, the International Federation of Clinical Chemistry and Laboratory Medicine Working Group on CGM (IFCC WG-CGM) has proposed a Dynamic Glucose Regions (DGR) framework that maps glucose concentration and rate of change jointly into five risk regions (BG Low, BG High, Alert Low, Alert High, Stable) and mandates that clinical trials sample a minimum proportion of points in each critical region rather than allowing recordings to be dominated by stable euglycemia.^30^ This guideline also proposes twelve explicit minimum-acceptance criteria spanning point accuracy, trend accuracy, and sensor-to-sensor consistency, evaluated separately in each dynamic region.^30^ Critics of the IFCC proposal have noted that some of its thresholds, for example, allowing up to 9% of paired points in the hypoglycemic range to exceed ±40% error, still permit clinically significant misses (a reading of 91 mg/dL when the true value is 65 mg/dL, falsely reassuring an automated insulin-delivery algorithm) and that confidence-bound-based criteria, as used in the FDA’s iCGM controls, would be more conservative than the point-estimate thresholds in the current IFCC draft^.29,30^

Taken together, these tools form a mature, increasingly sophisticated toolbox for subcutaneous CGM. None of them, however, was designed to answer the question that precedes all of them for an NI-CGM device under development: does the raw physical signal contain glucose-specific information at all, and is the test window long enough and variable enough to reveal whether it does?

## 3. Why Legacy Metrics Can Mislead When Applied to NI-CGM Algorithms

Four characteristics of NI-CGM development make the direct transfer of subcutaneous-CGM metrics problematic.

### 3.1 The signal itself is unproven

An electrochemical CGM sensor is built around a glucose oxidase reaction whose specificity for glucose is chemically established before any clinical evaluation begins; the evaluation question is how accurately and how stably that known reaction is transduced into a number. By contrast, non-invasive technologies rely on indirect approaches that imply complex measurement interpretation. For example, optical and photoacoustic NI-CGM signals are a composite of glucose absorption together with water content, temperature, perfusion, motion, ambient humidity, and sensor-skin contact quality.^32,33^ The raw photoacoustic amplitude is approximately proportional to optical power, the absorption coefficient, the photoacoustic cell pressure, and inversely related to cell volume and a 3/2 power of the modulation frequency. None of these terms are glucose-specific in isolation. Multiple authors developing mid-infrared photoacoustic NI-CGM have therefore built layered skin models (stratum corneum, granulosum, spinosum, dermis, blood compartment) and full digital twins of the optical, thermal, and acoustic chain specifically to separate glucose-related signal from these confounders before any prediction algorithm is trained.^32,34^ Practical experience with multi-sensor NI-CGM wearables in free-living conditions confirms this. Caduff et al. found subject-specific variability to be the dominant factor in glucose estimation error from a device combining dielectric spectroscopy, optical, temperature, humidity, and motion sensors. Their developed personalized model achieved MARD of 17.8% versus 21.1% for a population-level model variation driven largely by inter-individual differences in skin properties and sensor contact rather than by glucose-specific signal quality.^35^ Acciaroli et al., extending the same platform, showed that a calibration module anchored to an initial reference value contributed a larger performance improvement than any algorithmic refinement, underscoring how much of the apparent accuracy was attributable to the calibration offset rather than to a glucose-specific signal.^36^ If the evaluation pipeline jumps directly to MARD without first verifying that the signal carries information beyond these confounders, an apparently reasonable accuracy figure may reflect an algorithm fitting to skin temperature, motion, or calibration offset rather than to glycemia.

### 3.2 Real-world test windows are short and low-variability relative to legacy CGM validation studies

Clinical trials of NI-CGM devices remain few in number, free-living NI-CGM trials are scarcer still and less tightly controlled than the studies historically used to validate legacy subcutaneous CGM. A recent systematic review by Zhang et al. (2026) ^37^ quantified this systemic asymmetry, revealing that the median validation duration for NIGM studies is a mere 4 hours, compared to 14 days for established iCGMs, with 60% of NIGM cohorts conducting validation over 4 hours or less. In an eight-participant, free-living trial of a wearable QCL photoacoustic NI-CGM device, the average single recording (“experimentation”) lasted 5 hours and 40 minutes, and a test split using the final 30% of a recording therefore averaged only 1 hour 40 minutes. Over such a short window, especially in free-living, non-clamped conditions, the natural glycemic excursion of a participant is frequently smaller than either the sensor’s physical limit of detection or its intrinsic signal-to-noise ratio. Under these conditions, a trivial model that simply repeats the initial calibration value for the rest of the window can be very difficult to beat, not because it is accurate, but because there is little glycemic variation to predict.

Zhang et al. ^37^ quantified this duration-dependent bias systematically across the NI-CGM field: devices reported a pooled MARD of 8.7% in studies lasting four hours or less, but this figure degraded to 15.2% in studies exceeding 24 hours. Compounding this, 85% of NI-CGM studies omit performance testing in the hypoglycemic range (below 70 mg/dL) entirely, avoiding the glycemic excursions that would most clearly expose sensor limitations. Short and stable test windows therefore fail not only to capture long-term sensor drift and physiological confounds, but also to reveal whether a device can distinguish glycemia from the many non-glucose signals that dominate its output during periods of low variability, precisely the scenario in which an attractively low MARD can be reported with no true clinical meaning.

### 3.3 Reference-error propagation is amplified by the absence of an established NI-CGM accuracy floor

Traditional MARD calculations implicitly assume the comparator device is a perfect measure of true blood glucose; in practice, reference measurements carry their own error distribution which implies that the calculated value is relative to the control system and not absolute.^14^ For mature CGM, this error is a known, bounded nuisance, because the floor of achievable MARD with current subcutaneous sensors (approximately 7–9%) is well documented.^14,19^ For NI-CGM, no such consensus floor exists, and several authors have already observed that the literature reports MARD values of only a few percent for non-invasive devices without accompanying clinical evidence or any product reaching market, a pattern consistent with reference-error masking, short test windows, or both, rather than genuine sensor performance.^30^

### 3.4 Generalization level is rarely stated explicitly

A NI-CGM algorithm can be trained and tested at progressively broader levels of generalization: within a single recording session (intra-experimentation, capturing one specific skin/device contact configuration); across sessions from the same patient (intra-patient, capturing average skin and positioning variability for that individual); across sessions pooled from multiple patients where all participants appear in both training and test sets, split chronologically (intra-patient generalized, which exposes the algorithm to inter-individual skin diversity while still allowing the test participant’s physiology to be represented in training); or on patients entirely unseen during training (inter-patient, the most demanding test and the ultimate goal for a generalizable product). These levels are not interchangeable, and a MARD value reported without specifying which level was used cannot be compared to a value from another study, even for the same device. Section 4.5 defines each level and proposes that it should always be reported alongside any accuracy figure.

## 4. A Methodology for Demonstrating NI-CGM Glucose Specificity

We propose that the question of NI-CGM glucose specificity be addressed directly, prior to and alongside the clinical accuracy metrics described in Section 2, through the five elements summarized in Table 1 and detailed below. Three of these elements: the noise control, the constant-glycemia control, and the sensor-ablation control are negative controls in the strict experimental sense: each removes a candidate source of apparent accuracy (information content, time, and the candidate physical channel itself, respectively) and asks whether performance survives. A specificity claim is only as strong as the controls it survives. The methodology is intended for the device-development phase, to guide internal go/no-go decisions, and to make published accuracy claims interpretable. It is a deliberate complement to, not a substitute for, the clinical and regulatory metrics (ISO 15197 agreement rates, iCGM special controls, IFCC DGR criteria) required for market authorization. We return to that relationship in Section 6.1.

**Table 1.** Proposed five-elements methodology for demonstrating NI-CGM glucose specificity during algorithm development, intended to complement (not replace) ISO 15197 / iCGM / IFCC DGR clinical accuracy reporting.

| Step | Purpose | Operational description |
| --- | --- | --- |
| **1. Signal-quality gating** | Exclude unusable records before any algorithm training | Analysis of records for adequate signal-to-noise ratio and for adherence to the expected physical relationship (e.g., the $f^{-3/2}$ modulation-frequency dependence for photoacoustic signals); discard or flag records below a pre-defined coefficient-of-determination ($R^2$) threshold. |
| **2. Noise-control benchmark** | Test whether the recorded signal carries any glycemic information | Replace the physical signal recorded features with a centered normalized Gaussian white noise; train and test the candidate algorithm identically on this synthetic dataset. The algorithm must significantly outperform this noise floor on real sensor data, or hardware/feature-extraction must be revisited before further software optimization. |
| **3. Constant-glycemia benchmark** | Expose bias from short or low-variability test windows | Compare the candidate algorithm to a trivial model that simply propagates the initial calibration value forward through the test window. If this naive model is not beaten, either the test window is too short or the participant's glycemic excursion did not exceed the sensor's limit of detection. |
| **4. Sensor-ablation benchmark** | Isolate the contribution of the candidate physical signal from auxiliary covariates | Re-train and re-test the identical algorithm and pipeline with the candidate physical channel (e.g., the mid-infrared photoacoustic signal) removed, retaining only auxiliary covariates (motion, skin temperature, PPG). A material degradation in accuracy when the candidate channel is removed is direct, internal evidence that this channel, not only the auxiliary sensors, carries glucose-specific information. |
| **5. Explicit generalization level** | Make accuracy figures comparable across studies | Report whether train/test splitting was intra-experimentation, intra-patient, intra-patient generalized, or inter-patient (Table 2), with non-overlapping, chronologically ordered splits; never randomly shuffled splits for time-series data. |

### 4.1 Step 1 Signal-quality gating before algorithm training

Because the NI-CGM candidate signal is a composite of glucose-specific and non-specific contributions, the first evaluation step should occur before any predictive algorithm is trained: a check that each recorded data segment is itself usable. Two complementary checks have been described for photoacoustic NI-CGM and generalize to other optical and impedance-based modalities: (i) a signal-to-noise check, comparing the absolute signal amplitude with the device worn against the amplitude with the device non-coupled to skin, with a typical acceptance criterion of a factor three;^13^ and (ii) an internal consistency check, exploiting any known physical relationship within the signal, for instance, the theoretical f⁻³^⁄^² dependence of photoacoustic amplitude on modulation frequency and discarding records whose measured relationship departs from the expected one beyond a chosen coefficient-of-determination (R²) threshold. Reporting this gating step, and the fraction of data excluded by it, allows readers to judge how much of a reported accuracy figure rests on a pre-filtered, best-case subset of the recording.

### 4.2 Step 2 The noise-control benchmark

The purpose of this control is to answer, before any tuning of model complexity, whether the candidate algorithm’s apparent performance reflects genuine glycemic information in the signal or merely the algorithm’s capacity to fit any well-behaved numerical target. The procedure replaces the physical signal recorded features with synthetic centered normalized Gaussian white noise.

The candidate algorithm is evaluated on this synthetic dataset using an identical pipeline to the one used on real sensor data. If the algorithm’s performance on real signals is not statistically superior to its performance on this noise floor, the implication is that the physical sensor has not captured information relevant to physiology, and that further algorithmic refinement will not solve a hardware sensitivity problem.

### 4.3 Step 3 The constant-glycemia benchmark

The constant-glycemia benchmark assumes blood glucose remains entirely flat for the duration of the test window, propagating only the initial calibration value, as the prediction for every subsequent time point. This trivial model is mathematically simple but can be very difficult for sophisticated algorithms to outperform whenever the test window is short or the participant’s true glycemic excursion is small relative to the sensor’s physical limit of detection. In free-living, non-clamped conditions, which by design better reflect real-world device use than controlled clamp studies, but in which large or fast glycemic swings are not deliberately induced, this benchmark is a realistic and informative comparator rather than a strawman.

### 4.4 Step 4 The sensor-ablation benchmark and explainability

To isolate the contribution of the candidate channel, the sensor-ablation benchmark re-trains and re-tests the identical algorithm and pipeline with the candidate physical channel removed from the feature set, while every auxiliary covariate is retained unchanged. This approach is formally equivalent to feature ablation in the explainability literature, a method recognized for its high faithfulness, meaning it accurately reflects the true decision boundaries of the model rather than an approximation, on complex tabular data. ^38^

In the sensor-ablation benchmark, if the accuracy is materially worse without the candidate channel than with it, the channel is contributing information the auxiliary covariates cannot supply on their own, direct, internal evidence of specificity. If accuracy is unchanged or improves when the candidate channel is removed, the apparent performance of the full system was being carried by the auxiliary covariates (or by the benchmarks of Steps 2 and 3) The candidate physical signal has not yet been shown to be glucose-specific, regardless of how favorable the full-system MARD appears.

Beyond strict ablation, this step can be complemented by local explainability tools such as SHAP (SHapley Additive exPlanations) values ^38^. By assigning an importance score to each feature based on its marginal contribution to the model’s predictions, SHAP values continuously verify that the algorithm relies on the physical channel rather than predominantly on auxiliary proxies. It provides an additional, ongoing check on specificity as algorithm development progresses. Together, feature ablation and SHAP-based attribution form the most direct tests available, at the algorithm-development stage, for answering the question motivating this article: is the device measuring glucose? We report both in the case study below (Section 5.3).

The noise-control and constant-glycemia benchmarks (Steps 2 and 3) establish whether the recorded data, taken as a whole, carry exploitable information beyond chance and beyond the passage of time. Neither, by itself, identifies which channel within a multi-sensor NI-CGM device is responsible for that information. Most NI-CGM systems integrate the candidate physical signal (optical, photoacoustic, bioimpedance, Raman spectroscopy or microwave) alongside auxiliary covariates collected for motion compensation, calibration, or context: accelerometry, skin temperature and hygrometry, ambient conditions, and photoplethysmography (PPG).^13^ Several of these auxiliary signals correlate with glycemia indirectly, through autonomic and circulatory physiology. As a consequence, a model that performs well with the full sensor suite has not, by that fact alone, demonstrated that the candidate physical channel itself is glucose-specific.

The sensor-ablation benchmark isolates this question directly: the candidate algorithm and pipeline are re-trained and re-tested with the candidate physical channel removed from the feature set, while every auxiliary covariate is retained unchanged. If accuracy is materially worse without the candidate channel than with it, then the channel is contributing information the auxiliary covariates cannot supply on their own direct, internal evidence of specificity. If accuracy is unchanged or improves when the candidate channel is removed, the apparent performance of the full system was being carried by the auxiliary covariates (or by the benchmarks of Steps 2 and 3), and the candidate physical signal has not yet been shown to be glucose-specific, regardless of how favorable the full-system MARD appears. This control is the most direct test available at the algorithm-development stage of the question motivating this article is the device measuring glucose, and we report it explicitly in the case study below (Section 5.3).

### 4.5 Step 5 Explicit reporting of the generalization level

Time-series data must be split into training and test sets without temporal overlap; random shuffling, common in non time-dependent machine-learning problems, is inappropriate for physiological time series and can produce artificially optimistic accuracy estimates. Beyond this baseline requirement, NI-CGM developers can choose among four levels of generalization, each answering a different question and each yielding a numerically different MARD for the same underlying device (Table 2).

**Table 2.** Four levels of train/test generalization for NI-CGM algorithm development. A reported MARD is only interpretable once the generalization level is specified.

| Generalization level | Definition | Advantage / limitation |
| --- | --- | --- |
| **Intra-experimentation** | A separate algorithm is trained per recording session, using the first ~70% of records to train and the last ~30% to test. | Captures the specific skin/device contact of that session; limited by the small data volume available per session. |
| **Intra-patient** | 70% of a given patient's recording sessions are used for training, the remaining 30% of that same patient's sessions for testing. The device-skin configuration seen during testing has therefore been encountered during training. | Captures average skin configuration for one individual with more training data. Cannot test generalization to a new patient's physiology or device positioning. |
| **Intra-patient generalized** | All patients' experimentation contributes to both training and test sets, but test sessions are held out chronologically. Because each test participant is also present in the training set, inter-individual skin variability is represented in training, unlike strict intra-patient. | More training data and greater inter-individual physiological diversity than strict intra-patient. Sits between intra-patient and inter-patient in its test of generalization, since the test participant's own data appears in training. |
| **Inter-patient** | Training set is done on a subset of patients and testing blindly on entirely unseen patients. | Captures the highest physiological variability, it is the most demanding test of generalization and the most sensitive to inter-device variability. |

## 5. Illustrative Case Study: Neogly™ and the SKAMo-2 Trial

To illustrate how the proposed specificity-demonstration methodology changes the interpretation of a real NI-CGM dataset, we summarize results from Neogly™ (Eclypia, Grenoble, France), a body-worn NI-CGM combining multiple mid-infrared quantum cascade lasers (QCL) with a photoacoustic detection cell, evaluated in the SKAMo-2 clinical trial.^13^ The Table 3, the datasheet of Neogly^TM^ provide the main feature and physical signals recorded during the clinical trial SKAMo-2.

**Table 3.** Neogly datasheet.

| Characteristic | Specification |
| --- | --- |
| Dimensions | Measurement Head : 70*38*12 mm; Power Box : 100*65*20 mm |
| Total weight | 236 g |
| wearing | flexible wristband (In house) |
| HMI | Android App (In house) |
| Data storage | Embedded and cloud (AWS) |
| Communications | Bluetooth low energy, GSM (ST, BLUENRG) |
| Battery capacity | 2500 mAh (Avg : 200 mA) |
| Autonomy | 12 h (1 battery) |
| Electronics | 3 PCB (In house design, Eolane) |
| Main modality | Photoacoustic MIR (In house) |
| Laser(s) QCL | Pulsed, adjustable DC and frequency (Alpes Laser) |
| Optical power, beam combiner | 5 mW (In house design, Femtoprint) |
| Complementary modalities | PPG, accelerometer, microphone, temperature and hygrométrie (Different suppliers) |
| Control | Microcontroller (STM32L496QGI6P) |
| Data | Digital, 16 bits, 48 kHz (In house firmware) |
| EN ISO compliance | 10993-1, 13485, 14971, 60601-1, -2, -8, -11, 60825-1 (Dedicated tests) |

SKAMo-2 was a prospective, monocentric, free-living trial conducted at Grenoble Alpes University Hospital implying eight participants with type 1 diabetes, aged 18–50 years, registered with EudraCT 2022-A02789-34 and ClinicalTrials.gov NCT06035367. The device was worn continuously (day and night) for 7 to 9 days while participants continued their usual diabetes self-management, including their own subcutaneous CGM, which served as the reference for the prediction algorithm. The trial’s primary endpoints were technical feasibility and patient safety, with glycemia-prediction performance treated as a secondary, exploratory objective.^13^

We precise that our dataset is not large enough to extract MARD uncertainties estimations while building a model. It is an issue as we precisely aim at comparing MARD values, and we strongly recommend, if possible, to compute it. Our main proxy about the MARD variability is displayed in Fig. 1, as we represent how the MARD improves while injecting more and more data. We observe variations around an overall trend that amplitude is about 1%.

**Fig 1:**
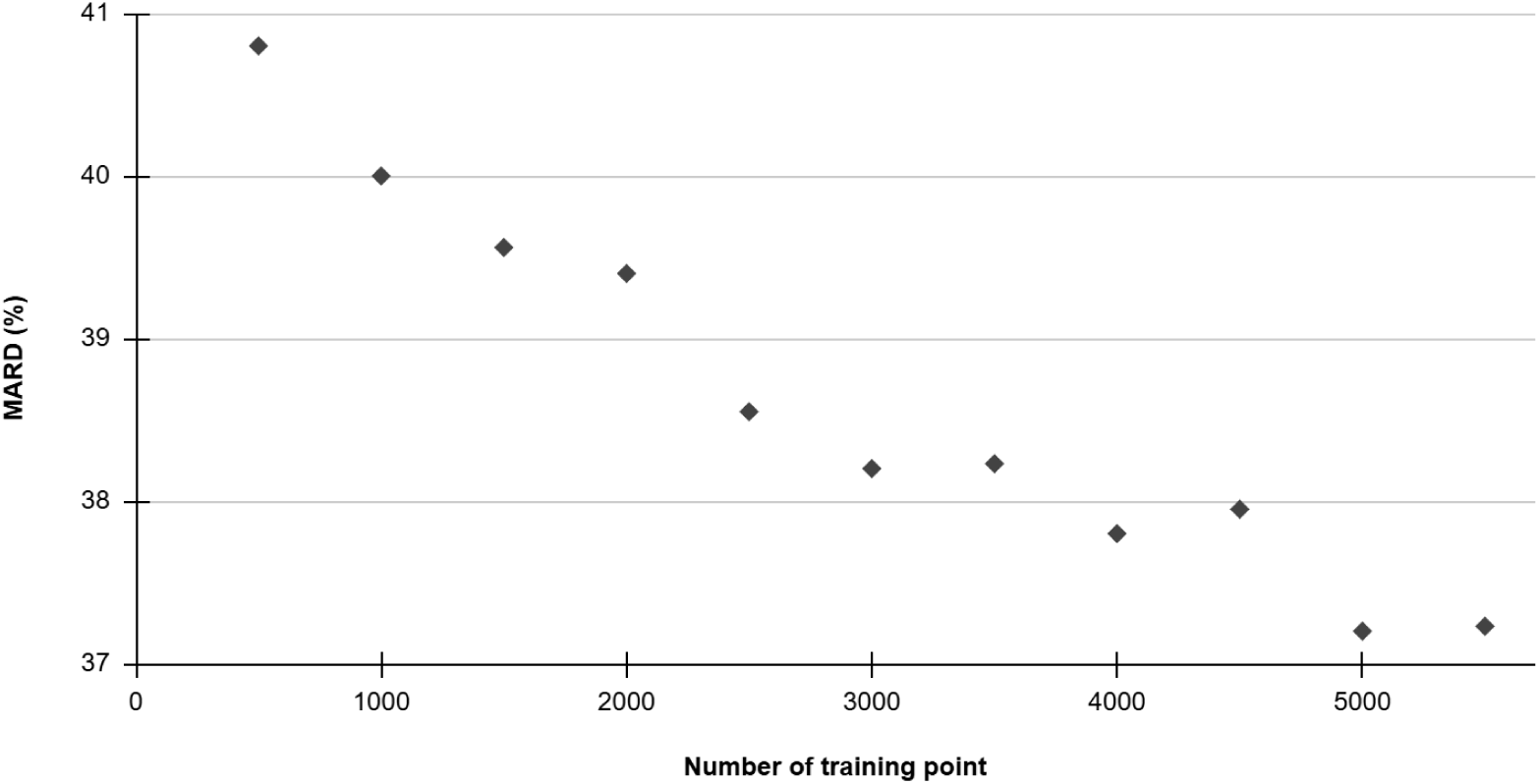
MARD in function of the number of data point in the training set

### 5.1 Step 1 in practice: data volume and signal-quality gating

Across the eight participants, 172 recording sessions longer than one hour were collected; 24 were excluded for system error or poor wrist positioning, leaving 148 usable sessions comprising 18,936 individual records and more than 750 hours of recording time (average session length: 5 hours 40 minutes). Device-wear adherence, measured as the percentage of total trial time the device was worn, ranged from 33% to 81% across participants, corresponding to more than 10 hours per day worn for seven of the eight participants, evidence of good adherence to a free-living wearable protocol. Applying the R²-based outlier criterion described in Section 4.1, with a threshold of 0.90 chosen as the inflection point (“elbow”) of the trade-off between record integrity and remaining data volume, excluded a further 24% of records, leaving 14,352 records as the final dataset used for algorithm development. In a related evaluation of the same device, the signal-to-noise ratio (worn versus unworn amplitude) exceeded the pre-specified threshold across 100% of sessions once outliers had been removed confirming that the gating step successfully identifies records where the device was not in adequate skin contact rather than simply discarding informative data.

### 5.2 Steps 2, 3, and 5 in practice: benchmarked MARD across generalization levels

Four algorithms of increasing complexity: linear regression, random forest, gradient-boosted random forest, and a dense neural network, were trained and benchmarked against the noise-control and constant-glycemia controls described in Sections 4.2 and 4.3, at each of the four generalization levels described in Section 4.5. The features used as input of algorithms are PA signals and complementary modalities presented in Table 3. The explicit true glycemia value was supplied to the algorithm as a calibration point at the start of each test segment, as is standard practice for factory-calibrated CGM evaluation. Results are summarized in Table 4 and present the algorithm with the best performances. Due to the very low reproducibility among devices during the clinical trial, the inter-patient level is not presented here. The sensor-ablation control (Step 4, Section 4.4) is reported separately in Section 5.3.

**Table 4.** MARD (%) for the noise-control benchmark, the constant-glycemia benchmark, and the best-performing machine-learning algorithm, across the three first generalization levels, for the Neogly™ NI-CGM device in the SKAMo-2 trial.

| Generalization level | Noisy-CGM control MARD (%) | Constant-glycemia control MARD (%) | Best ML algorithm | Best ML MARD (%) |
| --- | --- | --- | --- | --- |
| **Intra-experimentation** | 25.0 | 17.2 | Random forest | 22.5 |
| **Intra-patient** | 48.0 | 55.0 | Random forest | 39.0 |
| **Intra-patient generalized** | 52.0 | 55.0 | Dense neural network | 35.0 |

Three observations follow directly from reading these results as part of a specificity-demonstration methodology, rather than as an isolated accuracy claim.

First, at the intra-experimentation level, every algorithm outperformed the noise-control benchmark (25.0%), indicating that the recorded photoacoustic signal does carry physiological-relevant information; this is the central, necessary condition established by Step 2 of the methodology. In addition an algorithm trained on pure white noise achieved a MARD of 25% over an average test window of approximately 100 minutes, a value that would be considered, in isolation and by naive comparison to legacy CGM thresholds, surprisingly close to clinically usable. This result by itself demonstrates why MARD cannot be interpreted without a noise-floor comparison for NI-CGM.

Second, at this same level, the constant-glycemia benchmark (17.2%) outperformed the best machine-learning model (22.5%): because the test segment in this configuration averages only 1 hour 40 minutes in free-living, non-clamped conditions, the participant’s true glycemic variation was frequently smaller than the device’s effective limit of detection, exactly the scenario Step 3 of the methodology is designed to expose. Again this value of 17.2% would appear acceptable by historical CGM standards if reported alone, despite representing no predictive capability whatsoever.This demonstrates the need of benchmarking.

Third, moving to the intra-patient and intra-patient generalized levels reverses this relationship: the best algorithm (random forest, then a dense neural network) outperformed both controls, and absolute MARD improved as the training set grew from approximately 4 hours of training data per algorithm at the intra-experimentation level, to several hundred to several thousand records as data were pooled across sessions and patients. In the Figure 1 we verified this pattern directly by training intra-patient generalized models on progressively larger subsets of the pooled dataset and observing a corresponding, near-asymptotic improvement in MARD as the training set grew toward roughly 5,000 points, evidence that the signal contains exploitable, generalizable information rather than session-specific artifact.

### 5.3 Step 4 in practice: the sensor-ablation benchmark and direct evidence of specificity

The results in Section 5.2 establish that the full sensor suites the mid-infrared photoacoustic (PA) signal together with accelerometry, skin temperature/hygrometry, ambient conditions, and PPG carries exploitable glycemic information once enough data are pooled across sessions and patients. They do not, by themselves, establish that the PA channel is responsible for that information rather than the auxiliary covariates. We addressed this directly with the sensor-ablation benchmark described in Section 4.4: the identical algorithm and pipeline were re-trained and re-tested with the Mid-IR PA signal removed from the feature set, retaining accelerometry, skin temperature/hygrometry, and PPG unchanged, at both the intra-experimentation and inter-patient generalization levels.

Removing the Mid-IR PA signal degraded performance at both levels (Table 5). At the intra-experimentation level, MARD rose from 22.5% with the PA signal to 23.9% without it (RMSE 53.7 mg/dL versus 58 mg/dL). At the generalized intra-patient level, where the largest and most physiologically variable dataset is used, the effect was substantially larger: MARD rose from 35.0% with the PA signal to 43.1% without it (RMSE 65 mg/dL versus 77.6 mg/dL). The generalized intra-patient degradation is the most informative of the two results as it is the most demanding test of generalization and the least likely to be explained by a single session’s idiosyncratic calibration or contact configuration. As a consequence, this test is the most sensitive to the PA channel removal.

**Table 5.** Device-plus-algorithm MARD and RMSE with and without the Mid-IR photoacoustic (PA) signal, at the intra-experimentation and inter-patient generalization levels, for the Neogly™ NI-CGM device in the SKAMo-2 trial. Auxiliary covariates (accelerometry, skin temperature/hygrometry, PPG) are retained in both conditions. Data from Coutard et al.^13^

| Generalization level | MARD with Mid-IR PA (%) | MARD without Mid-IR PA (%) | RMSE with PA (mg/dL) | RMSE without PA (mg/dL) |
| --- | --- | --- | --- | --- |
| **Intra-experimentation** | 22.5 | 23.9 | 53.7 | 58 |
| **Intra-patient generalized** | 35.0 | 43.1 | 65 | 77.6 |

This is, to our knowledge, the most direct evidence available from this case study that the candidate physical channel, not only the auxiliary motion, temperature, and PPG covariates retained in the ablated condition, is contributing to glucose-specific information. The trend observed with the MARD is confirmed by the significant decrease in the root mean square error (RMSE), which is another statistical tool that can help overcome certain limitations of the MARD. These elements do not, on its own, establish clinical accuracy: the absolute MARD values in both conditions remain far from competitive with subcutaneous CGM (Section 5.2). The result solves the specificity question rather than the accuracy one. However, it directly answers the interrogation that motivates this article, in a way no aggregate MARD value computed on the full sensor suite alone can: removing the PA channel costs accuracy, at every generalization level tested. It is the operational definition of specificity we use throughout this article.

None of the absolute MARD values reported here (37–39% for the best inter- and intra-patient models) would be considered competitive with current-generation, factory-calibrated subcutaneous CGM systems, which MARD against laboratory reference methods is generally reported in the single digits to low tens.^2,3^ It is explicit that this is a preliminary, feasibility-stage result rather than a claim of clinical performance, and that signal stability, reproducibility, and removal of glucose-unrelated physiological variation remain the principal areas for further development. What the case study illustrates is methodological rather than competitive. Reporting these MARD values without the accompanying noise, constant-glycemia, and sensor-ablation benchmarks, and without specifying the generalization level, would have allowed the intra-experimentation figure (18.3%) to be read, by analogy with subcutaneous CGM literature, as an encouraging accuracy result. But in fact it was statistically indistinguishable from a model with zero predictive content over that specific, short test window and would have left the device’s central claim, that it measures glucose specifically, entirely unsupported by the kind of internal control that Table 4 provides.

## 6. Discussion

### 6.1 Relationship to existing scoping reviews and consensus efforts

The proposal made here is consistent with, and builds on, two existing strands of work. A 2023 scoping review of 129 CGM clinical performance studies published between 2002 and 2022 documented wide heterogeneity in population size (6 to 318 subjects), comparator sample type, reference device, and pairing method, concluding that this heterogeneity undermines direct comparison of reported MARD values even within the mature, subcutaneous CGM literature, and issued detailed reporting recommendations.^11^ Separately, the IFCC WG-CGM’s proposed Dynamic Glucose Regions framework directly addresses the static, point-in-time limitation of MARD and the Clarke/Parkes grids by requiring a minimum proportion of test data in clinically critical, dynamic glucose regions.^28^ Both efforts target the clinical-trial design and reporting stage, generally assuming that a working, calibrated sensor and algorithm are already available, and that the question being asked is how accurate that sensor is across the glycemic range. The methodology proposed in the present article asks a different, prior question, and is therefore intended to sit upstream of these efforts rather than in competition with them: during algorithm development, before the IFCC DGR question of range-stratified accuracy can be meaningfully posed, the present methodology asks whether a usable, glucose-specific signal exists at all. We see this as a deliberate division of labor rather than a gap in the IFCC proposal: IFCC DGR was not designed to answer the specificity question, and we do not think it should be redesigned to do so; instead, we propose that specificity be demonstrated first, using the controls in Table 1, and that IFCC DGR (or an equivalent clinical-accuracy framework) then be applied once that demonstration is complete. We see no tension between the two and recommend that both be required, at their respective stages, for any NI-CGM device progressing toward market.

### 6.2 Implications for regulatory and publication practice

We make four practical recommendations consistent with the methodology in Table 1. First, any published or submitted MARD, agreement-rate, or error-grid result for an NI-CGM algorithm should be accompanied by the noise-control and constant-glycemia benchmark values computed on the identical dataset and pipeline, so that readers can judge whether the reported accuracy reflects genuine signal content rather than test-window characteristics. Second, where the device combines a candidate physical signal with auxiliary covariates (motion, temperature, PPG, or similar), a sensor-ablation result should be reported showing accuracy with and without the candidate channel, so that a specificity claim rests on direct, internal evidence rather than on the performance of the full system alone. Third, the generalization level (intra-experimentation, intra-patient, intra-patient generalized, or inter-patient) should be stated explicitly in any abstract or summary table, not only in methods text, given how substantially it changes the resulting MARD (Table 4) and the apparent size of the sensor-ablation effect (Table 5). Fourth, signal-quality gating criteria and the fraction of data excluded by them should be reported, since accuracy figures computed only on a pre-filtered, best-case subset of recordings will not generalize to unselected, real-world use.^28^

These recommendations do not conflict with, and are intended to be used alongside, the formal clinical accuracy requirements that govern regulatory clearance: ISO 15197:2013 agreement-rate criteria, the FDA’s iCGM special controls under 21 CFR 862.1355, and the IFCC WG-CGM’s proposed DGR-based acceptance criteria.^18,20,21,29^ Rather, they address an earlier and narrower question whether a candidate physical signal, and the algorithm trained on it, has demonstrated genuine and generalizable glucose specificity that determines whether a device is ready to proceed to the larger, more expensive, range-stratified trials those regulatory frameworks require.

### 6.3 Limitations

This proposal has several limitations. The illustrative case study is drawn from a single candidate signal (mid-infrared photoacoustic spectroscopy) and a small participant cohort (eight individuals with type 1 diabetes), and the absolute MARD values reported should not be generalized to other NI-CGM technologies (bioimpedance, microwave, Raman spectroscopy, PPG-based machine learning, near-infrared spectroscopy) without modality-specific adaptation of the signal-quality gating and sensor-ablation steps.^5,7,35^ The noise-control, constant-glycemia, and sensor-ablation benchmarks, while informative, are necessary rather than sufficient conditions for clinical validity: beating the noise and constant-glycemia controls, and showing degraded performance under sensor ablation, demonstrates that an algorithm has learned something beyond noise, beyond a flat prediction, and beyond the auxiliary covariates alone, but does not by itself establish clinical accuracy in the sense required by ISO 15197 or iCGM criteria, which remain the appropriate standards for that purpose. The sensor-ablation control is also imperfect as an isolation technique: auxiliary covariates such as skin temperature or PPG-derived heart-rate variability can themselves correlate with glycemia through autonomic physiology, so a degradation in accuracy when the candidate channel is removed is strong evidence of specificity but does not, on its own, fully exclude every indirect physiological pathway. To evaluate the exact contribution of auxiliary sensors, the sensor-ablation should be repeated over these signals.

## 7. Conclusion

Every team developing a non-invasive glucose sensor eventually confronts the same question, and the answer cannot be assumed: is the device actually measuring glucose? Non-invasive CGM technologies are evaluated today with metrics built for a different problem, confirming the accuracy of a known, specific physiological signal rather than for establishing that a glycemia-specific signal exists in the first place and can be generalized across patients and conditions. Applied without modification, MARD and the Clarke/Parkes error grids can reward algorithms with no genuine predictive content, particularly over the short, low-variability test windows typical of early-stage, free-living NI-CGM trials. On the contrary, they can also draw negative conclusions about promising, under development, technologies. We have proposed a methodology: signal-quality gating, a noise-control benchmark, a constant-glycemia benchmark, a sensor-ablation benchmark, and an explicit reporting of the train/test generalization levels. The methodology is designed specifically to demonstrate glucose specificity, and to make NI-CGM accuracy claims interpretable and comparable across studies. We illustrated its use with a quantum-cascade-laser photoacoustic device evaluated in the SKAMo-2 free-living clinical trial, where the sensor-ablation results provided direct, internal evidence that the photoacoustic channel itself carries glucose-specific information beyond what auxiliary motion and temperature sensors supply on their own. This methodology is deliberately complementary to, and not a substitute for, established clinical accuracy frameworks such as IFCC WG-CGM’s Dynamic Glucose Regions guideline^28^: we see specificity demonstration and clinical accuracy characterization as two distinct, sequential questions, and recommend that the former be answered, explicitly and with the controls proposed here, before the latter is attempted. We encourage NI-CGM developers, reviewers, and journals to require these specificity benchmarks alongside conventional CGM accuracy metrics, as a precondition for interpreting any reported MARD, agreement rate, or error-grid result for a device of this class.

## Data Availability

The datasets generated and analyzed during the current study are available from the corresponding author upon reasonable request

## Acknowledgments

The authors would like to thank all individuals who contributed to this work. Special thanks go to the Eclypia team for their exceptional collaboration, commitment, and assistance at various stages of this development.

## Author Contributions

All persons who meet authorship criteria are listed as authors. All authors have participated sufficiently in the work to take public responsibility for the content, including participation in the concept,design analysis, writing or revision of the manuscript.

## Declaration of Conflicting Interests

The author(s) declared the following potential conflicts of interest with respect to the research, authorship, and/or publication of this article: Jean-Guillaume Coutard was the chief technology officer of Eclypia, Hélène Marie was the General Manager of Eclypia, Romain Blanc was the Data product manager, Pierre Yves Benhamou was in charge of the clinical investigation.

## Funding

This work was funded by ECLYPIA, France 2030 – PSPC (DIASBE program)

## Notes

### Clinical Trial

NCT06035367

### Author Declarations

The Ethics committee/IRB of Comitee de Protection des Personnes Ouest I gave ethical approval for this work (EudraCT 2022-A02789-34, ClinicalTrials.gov NCT06035367). Written informed consent was obtained from all participants prior to study inclusion.

